# The relationships between women’s reproductive factors: a Mendelian randomization analysis

**DOI:** 10.1101/2021.09.29.21264251

**Authors:** Claire Prince, Gemma C Sharp, Laura D Howe, Abigail Fraser, Rebecca C Richmond

## Abstract

**Background:** Women’s reproductive factors include their age at menarche and menopause, the age at which they start and stop having children, and the number of children they have. Studies that have linked these factors with disease risk have largely investigated individual reproductive factors and have not considered the genetic correlation and total interplay that may occur between them. This study aimed to investigate the nature of the relationships between eight female reproductive factors.

**Methods:** We used data from the UK Biobank and genetic consortia with data available for the following reproductive factors: age at menarche, age at menopause, age at first birth, age at last birth, number of births, being parous, age at first sex and lifetime number of sexual partners. Linkage disequilibrium score regression (LDSC) was performed to investigate the genetic correlation between reproductive factors. We then applied Mendelian randomization (MR) methods to estimate the causal relationships between these factors. Sensitivity analyses were used to investigate directionality of the effects, test for evidence of pleiotropy and account for sample overlap.

**Results:** LDSC indicated that most reproductive factors are genetically correlated (r_g_ range: |0.06 – 0.94|), though there was little evidence for genetic correlations between lifetime number of sexual partners and age at last birth, number of births and ever being parous (r_g_ < 0.01). MR revealed potential causal relationships between many reproductive factors, including later age at menarche (1 SD increase) leading to a later age at first sexual intercourse (Beta (B)=0.09 SD, 95% confidence intervals (CI)=0.06,0.11), age at first birth (B=0.07 SD, CI=0.04,0.10), age at last birth (B=0.06 SD, CI=0.04,0.09) and age at menopause (B=0.06 SD, CI=0.03,0.10). Later age at first birth was found to lead to a later age at menopause (B=0.21 SD, CI=0.13,0.29), age at last birth (B=0.72 SD, CI=0.67,0.77) and a lower number of births (B=-0.38 SD, CI=-0.44,-0.32).

**Conclusion:** This study presents evidence that women’s reproductive factors are genetically correlated and causally related. Future studies examining the health sequelae of reproductive factors should consider a woman’s entire reproductive history, including the causal interplay between reproductive factors.

## Introduction

A woman’s reproductive life course includes her age at menarche (AAM) and menopause, the age at which she starts and stops having children, and the number of children she has, as well as the age she first has sexual intercourse, and the number of sexual partners she has in her lifetime. Some of these reproductive factors have been identified as risk factors for chronic diseases, including breast cancer, respiratory disease and cardiometabolic diseases.^(1)^ A younger AAM and older age at menopause were associated with an increased risk of breast cancer in one large meta-analysis,^(2)^ while having fewer children and a higher age at first birth (AFB) were positively associated with breast cancer risk in another.^(3)^ Other studies have implicated AAM, AFB, number of still births and miscarriages, age at menopause and parity in relation to respiratory and cardiovascular disease.^(4-6)^ One study found that later AAM was associated with reduced risk of coronary artery disease.^(7)^ Having any children and later AFB have been associated with a lower risk of lung cancer.^(8)^ Older AAM and a shorter reproductive period have also been linked with higher risk of chronic kidney disease.^(9,10)^

However, on the whole, studies have not considered a woman’s entire reproductive history and the potential interplay between reproductive factors. Understanding the inter-relationships between reproductive factors is important to correctly identify potential confounders (common causes of the exposure and outcome of interest) and mediators (factors that lie on the causal pathway between exposure and outcome). Information on multiple reproductive factors will provide useful additions to algorithms for predicting disease risk in women.^(1)^

Evidence of association between AAM and menopause is inconsistent, with some studies reporting earlier AAM associated with earlier menopause,^(11-16)^ others showing the inverse association,^(17,18)^ and some showing no evidence of this association.^(19-24)^ While there is some evidence of an association between an earlier AAM and earlier AFB,^(25,26)^ there is little evidence of association between AAM and parity.^(26)^ Another study has also investigated reproductive factors in relation to sexual history, suggesting a younger AAM is not a risk factor for younger age at first having sexual intercourse (AFS).^(27)^ Associations between reproductive factors could be reflective of causal relationships, or common genetic or non-genetic environmental causes, i.e. confounding.

Observational studies are prone to confounding bias as it is difficult to capture all confounders accurately. Mendelian randomization (MR) is a method that assesses the causal relationship between an exposure and outcome by using genetic variants robustly associated with the exposure. MR is advantageous as it is less likely to be affected by confounding and reverse causation than standard multivariable regression analysis.^(28-30)^ There have been an increasing number of genome wide association studies (GWAS) of reproductive factors,^(31-33)^ which can be used to investigate genetic correlation (i.e. shared genetic causes) between these factors as well as whether relationships between reproductive factors may be causal using MR.

The present study aims to identify and clarify the nature of any relationships between women’s reproductive factors, by investigating their genetic overlap and the causal relationships between eight reproductive factors, including potential bidirectional effects where the temporal order between the traits is not clear.

## Methods

### UK Biobank

The UK Biobank study is a large population-based cohort of 502,682 individuals who were recruited at ages 37–73 years across the UK between 2006 and 2010. The study includes extensive health and lifestyle questionnaire data, physical measures, and biological samples from which genetic data has been generated. The study protocol is available online, and more details have been published elsewhere.^(34)^ At recruitment the participants gave informed consent to participate and be followed up.

### Reproductive factors

The reproductive factors investigated in this study were: age at menarche, age at menopause, age at first live birth, age at last live birth, number of live births, age first had sexual intercourse, lifetime number of sexual partners (at the time of assessment) and parous status (ever/never given birth at the time of assessment). In UK Biobank these reproductive factors were derived from questionnaire responses at the baseline assessment; further details can be found in the **Supplementary Note**.

#### Phenotypic correlation

We calculated the correlation between reproductive factors using the Pearson correlation coefficient.

### GWAS

To identify genetic variants robustly related to each of the reproductive factors, we first performed genome wide association studies (GWAS) for each reproductive factor among women in the entire UK Biobank sample. Each GWAS was performed using the Medical Research Council (MRC) Integrative Epidemiology Unit (IEU) UK Biobank GWAS pipeline.^(35,36)^ BOLT-LMM was used to conduct the analysis in the GWAS pipeline,^(37)^ which accounts for population stratification and relatedness using linear mixed modelling. Genotyping chip and age were included as covariates. Genome-wide significant SNPs were selected at p <5×10^−8^ and were clumped to ensure independence at linkage disequilibrium (LD) r^2^ < 0.001 and a distance of 10 000 kb using the TwoSampleMR package.^(36)^

### Genetic correlation

Genetic correlations between the reproductive factors were calculated using LDSC and the UK Biobank GWAS summary statistics.^(38,39)^ The regressions were performed using pre-computed LD scores for each SNP calculated based on individuals of European ancestry from using 1000 Genomes European data and are appropriate for use with European GWAS data.^(38)^ These LD scores were filtered to HapMap3 SNPs as these are well-imputed in most studies.^(40)^ SNPs found on chromosome 6 in the region 26MB to 34MB were excluded. GWAS summary statistics were converted for LDSC regression using the munge_sumstats.py command from the command line tool “ldsc”, and LDSC was performed using the ldsc.py command.

### Mendelian randomization

We conducted MR analysis using the “TwoSampleMR” R package,^(36)^ where the inverse variance weighted (IVW) method was used in the primary analysis to assess the causal relationships between pairs of reproductive factors. This method combines Wald ratios, calculated by dividing the SNP-outcome association by the SNP-exposure association, in a multiplicative random effect meta-analysis where the weight of each ratio is the inverse of the variance of the SNP-outcome association.^(41)^ We assessed earlier-occurring reproductive factors as the exposure in relation to later-occurring factors (the outcomes), e.g. AAM was investigated as a potential cause of age at menopause but not vice versa. In some cases where there was no clear temporal ordering, we carried out analyses in both possible directions, e.g., between ever parous status and age at menopause. These cases are shown in **Table S1**. All relationships tested by MR are shown in **Table S2**, and GWAS estimates were standardized (mean = 0 and standard deviation (SD) = 1) prior to performing MR.

The IVW method makes a number of assumptions: that the genetic instruments are strongly associated with the exposure; do not share common causes, either genetic or other confounders such as population stratification, with the outcome; and are not pleiotropic i.e., do not have an effect on the outcome through a pathway other than via the exposure. ^(41)^ We therefore performed a series of sensitivity analyses to evaluate the robustness of our results to these assumptions (see “Evaluating assumptions”).

In our primary analysis, we applied two-sample MR methods on a single large dataset, UK Biobank, which is advantageous over other methods due to the large sample size. In this analysis, the GWAS used for the exposure and outcome were both performed on women in the UK Biobank study, and therefore the exposure and outcome samples overlap entirely. Large overlap in the sample(s) used to generate genetic variant-exposure and genetic variant-outcome associations can introduce bias in estimates obtained using two-sample MR.^(42)^ In particular, sample overlap between the exposure and outcome samples may bias estimates towards the observational (and potentially confounded) exposure-outcome association and may lead to an overestimation of effects.^(42)^ While it has been proposed that this approach of applying two-sample MR methods in a single sample may be performed within large studies with minimal bias introduced to the causal estimates by sample overlap,^(43)^ we performed a series of sensitivity analyses to evaluate the robustness of our results to this (see “Assessing the impact of sample overlap”).

### Evaluating MR assumptions

We evaluated the likelihood that MR assumptions were violated where we found evidence of effects in our primary analysis.

#### Instrument strength

The strength of the genetic instrument for each reproductive factor in the main IVW analysis was assessed using the mean F statistic, calculated based on the variance explained (r^2^) by the genetic instrument and sample size of the exposure.^(30)^

#### Negative controls

We repeated our primary analysis for five ‘negative control’ pairs of reproductive factors, for which we would not expect to see causal effects due to their temporal ordering (the outcome occurring before the exposure). These negative controls included the effect of AFB on AAM, AFS on AAM, AFB on AFS, age at menopause on AFS, and ALB on AAM. In these cases, any evidence of an effect would suggest pleiotropy. This may occur when a genetic instrument affects the exposure and outcome through a shared heritable factor, which could be a shared process or pathway.^(44)^

#### Heterogeneity

We performed a test for heterogeneity using Cochran’s Q statistic using the TwoSampleMR package between instruments. A Q larger than the number of instruments minus one provides evidence for heterogeneity and invalid instruments, which can imply presence of pleiotropy.^(45,46)^

#### Pleiotropy

We used additional MR methods: Weighted mode,^(47)^ Weighted median,^(48)^ and MR Egger,^(49)^ to assess evidence of pleiotropy.^(50)^ The intercept and 95% confidence interval of the MR-Egger regression line was used to determine directional pleiotropy using the TwoSampleMR package.^(49)^ We also applied the R function MR-PRESSO (Mendelian Randomisation Pleiotropy RESidual Sum and Outlier) to identify and correct for potential outliers (p <0.05).^(51)^ Further details can be found in the **Supplementary Note**.

#### Steiger filtering for bidirectional relationships

We performed the MR Steiger test and Steiger filtering bi-directionally for the following pairs of reproductive factors where the temporal ordering was not clear.^(52)^ (**Table S3**) This was performed to assess whether the hypothesized causal directional of the relationship was correct for each genetic instrument.^(52)^ Further details can be found in the **Supplementary Note**.

### Assessing the impact of sample overlap

To investigate whether the degree of bias introduced by sample overlap impacted our findings, we conducted a series of sensitivity analyses.

Firstly, we performed MR on GWAS summary statistics using a ‘split-sample’ approach, in which the UK Biobank sample was divided in two halves at random. The MR analysis was performed twice for each relationship, once using the exposure GWAS from one half and the outcome GWAS from the second half and vice versa, with the resulting MR effect estimates being meta-analysed using a fixed effects model.

Secondly, we performed two-sample MR using results from largely non-UK Biobank replication studies and consortia to estimate SNP effects on the exposure (sample 1) and UK Biobank estimates to estimate SNP-effects on the outcome (sample 2), and vice versa where appropriate.^(31,32,53)^ Further details on the number of studies and sample sizes used for the replication consortia are shown in **Table S4**. Using replication studies may also avoid bias introduced by winner’s curse, which is the overestimation of SNP effects on the exposure in a discovery GWAS.^(54,55)^

Finally, we used a recently developed MR method, MRlap, that is robust to bias introduced by sample overlap, winner’s curse and weak instruments.^(56)^ MRlap was performed using the UK Biobank GWAS summary statistics for reproductive factors where both the exposure and outcome were continuous i.e., excluding associations involving ever parous status, as the correction for biases cannot account for a different degree of overlap for cases/controls in case of binary traits.^(56)^

Only those reproductive factor associations for which there was evidence of an effect from the primary analysis were taken forward for this sensitivity analysis, as the causal effect would likely be overestimated when performing MR with overlapping exposure and outcome samples.^(42)^

## Results

### UK Biobank

264 698 women from UK Biobank were included in this analysis. The mean age at assessment was 56.4 years (SD=8.0), further sample characteristics are shown in **Table 1**. Many of the reproductive factors were weakly phenotypically correlated. The strongest correlations were between AFB and ALB (Pearson correlation coefficient=0.71), and AFB and number of births (Pearson correlation coefficient=-0.34) (**Figure 1A**).

**Table 1.** UK Biobank reproductive factors descriptives.

| Reproductive factor |  |  |
| --- | --- | --- |
|  | N | Mean (SD) |
| Age at menarche (years) | 243 898 | 13.0 (1.6) |
| Age first had sexual intercourse (years) | 219 486 | 19.1 (3.6) |
| Age at first live birth (years) | 203 606 | 25.9 (5.1) |
| Age at last live birth (years) | 203 356 | 30.1 (5.2) |
| Age at menopause (years) | 143 791 | 49.7 (5.1) |
| Number of live births | 250 746 | 1.8 (1.2) |
| Reproductive factor | N | Median (IQR) |
| Lifetime number of sexual partners | 208 274 | 3 (4) |
| Reproductive factor | (% (N)) |  |
| Never parous | 18.69 (49 358) |  |

SD: standard deviation. IQR: Interquartile range. N: Sample size

**Figure 1.**
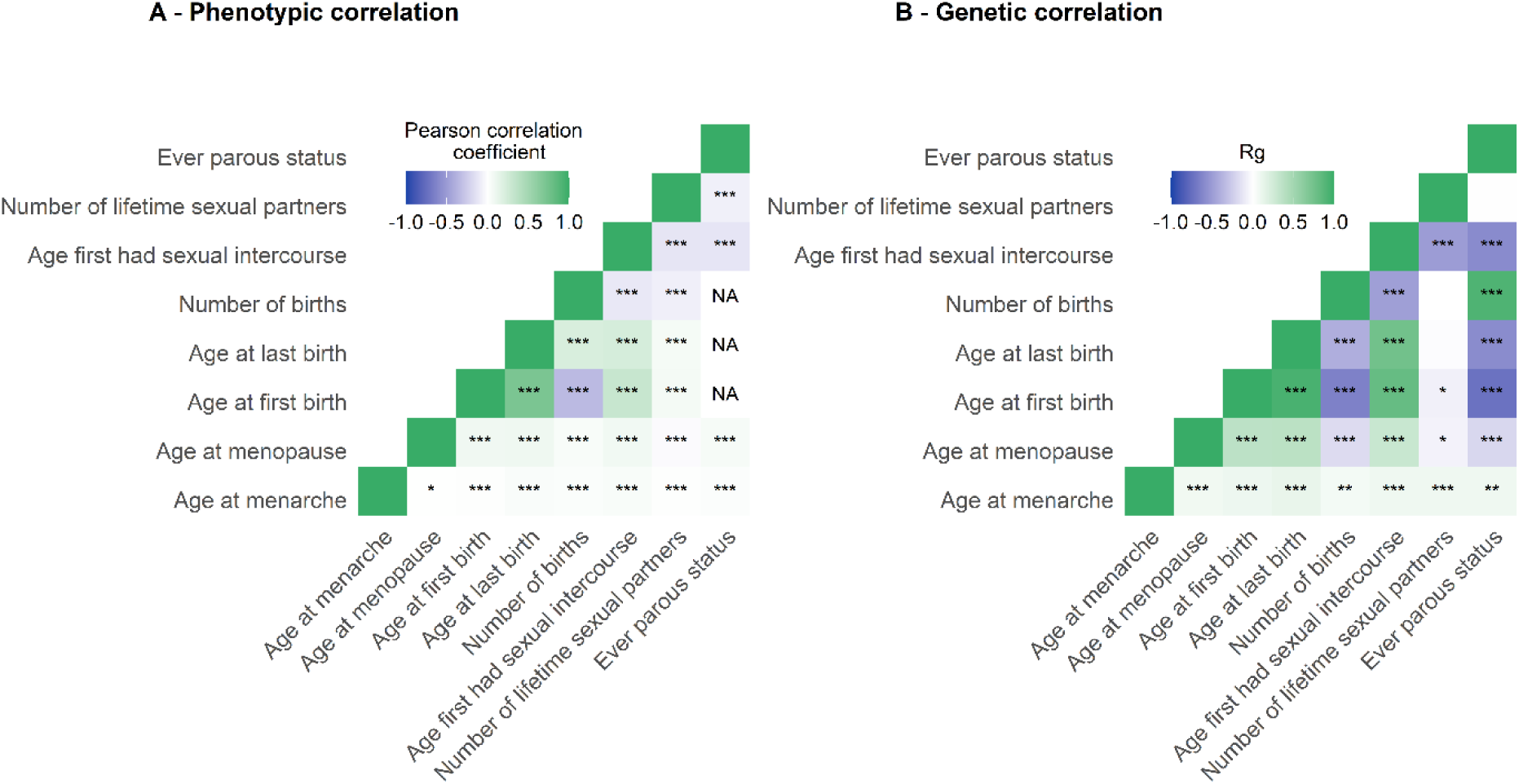
A - Phenotypic correlation using the Pearson correlation coefficient. B - Genetic correlation between reproductive factors using LD score regression. ***: <0.001; **: <0.01; *:<0.05.

### UK Biobank GWAS

**Table 2** displays the number of variants associated with the eight reproductive factors at genome-wide significance (p<5×10^−8^) after LD clumping within the full UK Biobank sample. Between four (ever parous status) and 223 (AAM) SNPs were identified. All F statistics were above the standard threshold of 10, indicative of strong genetic instruments (**Table 2**).

**Table 2.** Sample size of the exposure (N), F statistic and the number of SNPs (nSNPs) used within the primary analysis.

| Exposure | N | F statistic | R<sup>2</sup> | nSNPs |
| --- | --- | --- | --- | --- |
| Age at menarche | 243 898 | 74.95 (75.28 for outcome AFS) | 0.064 | 223 (222 for outcome AFS*) |
| Age at menopause | 143 791 | 85.08 | 0.047 | 84 |
| Age at first live birth | 203 606 | 41.97 | $8.38 \times 10^{-3}$ | 41 |
| Age at last live birth | 203 356 | 43.46 | $1.92 \times 10^{-3}$ | 9 |
| Number of live births | 250 746 | 46.75 | $1.68 \times 10^{-3}$ | 9 |
| Age first had sexual intercourse | 219 486 | 39.46 | $9.44 \times 10^{-3}$ | 53 |
| Lifetime number of sexual partners | 208 274 | 39.20 | $6.36 \times 10^{-3}$ | 34 |
| Ever parous status | 250 746 | 53.92 | $8.59 \times 10^{-4}$ | 4 |

AFS – Age first had sexual intercourse. \* One palindromic SNP was excluded from the MR between AAM and AFS.

### Genetic correlation

The LDSC revealed that the 8 reproductive factors were genetically correlated (r_g_ range: |0.06 – 0.94|), except for lifetime number of sexual partners, which was not correlated with ALB, number of births or ever parous status (r_g_ < 0.01). AAM was only weakly genetically correlated with other reproductive factors (r_g_ range: 0.06 - 0.12) In general, genetic correlations were larger in magnitude than the corresponding phenotypic correlations (**Figure 1B, Table S5**).

### Mendelian randomization

MR findings from the primary analysis suggest that the positive genetic correlation reflects a causal relationship between later AAM (1 SD increase) and later AFS (Beta (B)= 0.09, 95% confidence intervals (CI)= 0.06, 0.11), AFB (B=0.07 SD, CI=0.04, 0.10), ALB (B=0.06 SD, CI=0.04, 0.09) and age at menopause (B=0.06 SD, CI=0.03, 0.10) (**Figure 2A**). Findings suggest later AFB (1 SD increase) may lead to a later age at menopause (B=0.21 SD, CI=0.13, 0.29), later ALB (B=0.72 SD, CI=0.67, 0.77) and lower number of births (B=-0.38 SD, CI=-0.44, -0.32) (**Figure 2G**). In addition, later AFS (1 SD increase) appears to lead to later age at menopause (B=0.11 SD, CI=0.04, 0.18), later AFB (B=0.56 SD, CI=0.49, 0.63), later ALB (B=0.42 SD, CI=0.35, 0.50), lower number of births (B=-0.24 SD, CI=-0.31, - 0.17), lower lifetime number of sexual partners (B=-0.51 SD, CI=-0.58, -0.44) and increased likelihood of not having any children (OR=0.90 SD, CI=0.88, 0.93). Findings suggest later ALB (1 SD increase) may lead to a lower number of births (B=-0.19 SD, CI=-0.31, -0.07) (**Figure 2H**). Finally, a higher lifetime number of sexual partners decreases the likelihood of having children (OR=0.96 SD, CI=0.92, 1.0) (**Figure 2L**).

**Figure 2.**
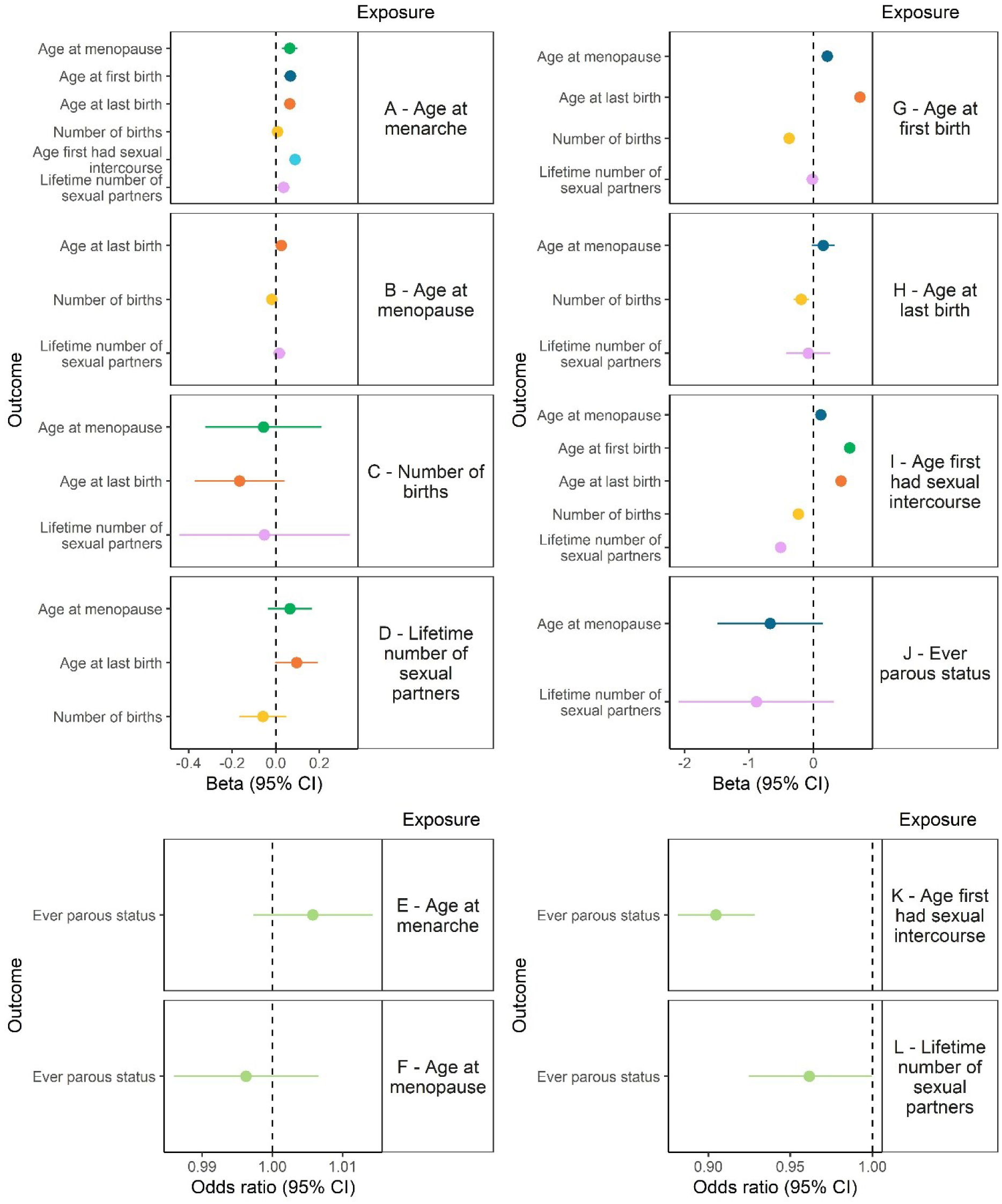
Relationships identified in the primary with evidence of a causal effect. + indicates a positive relationship and – indicates a negative relationship.

Age at menopause, number of births and ever having children do not appear to have strong effects on any of the other reproductive factors (**Figure 2B, C, F, J)**, although confidence intervals for the effects of number of births and ever having children are wide. Full results of this analysis can be found in **Table S6**, and a causal graph shows where we found evidence of an effect between reproductive factors (**Figure 3**).

**Figure 3.**
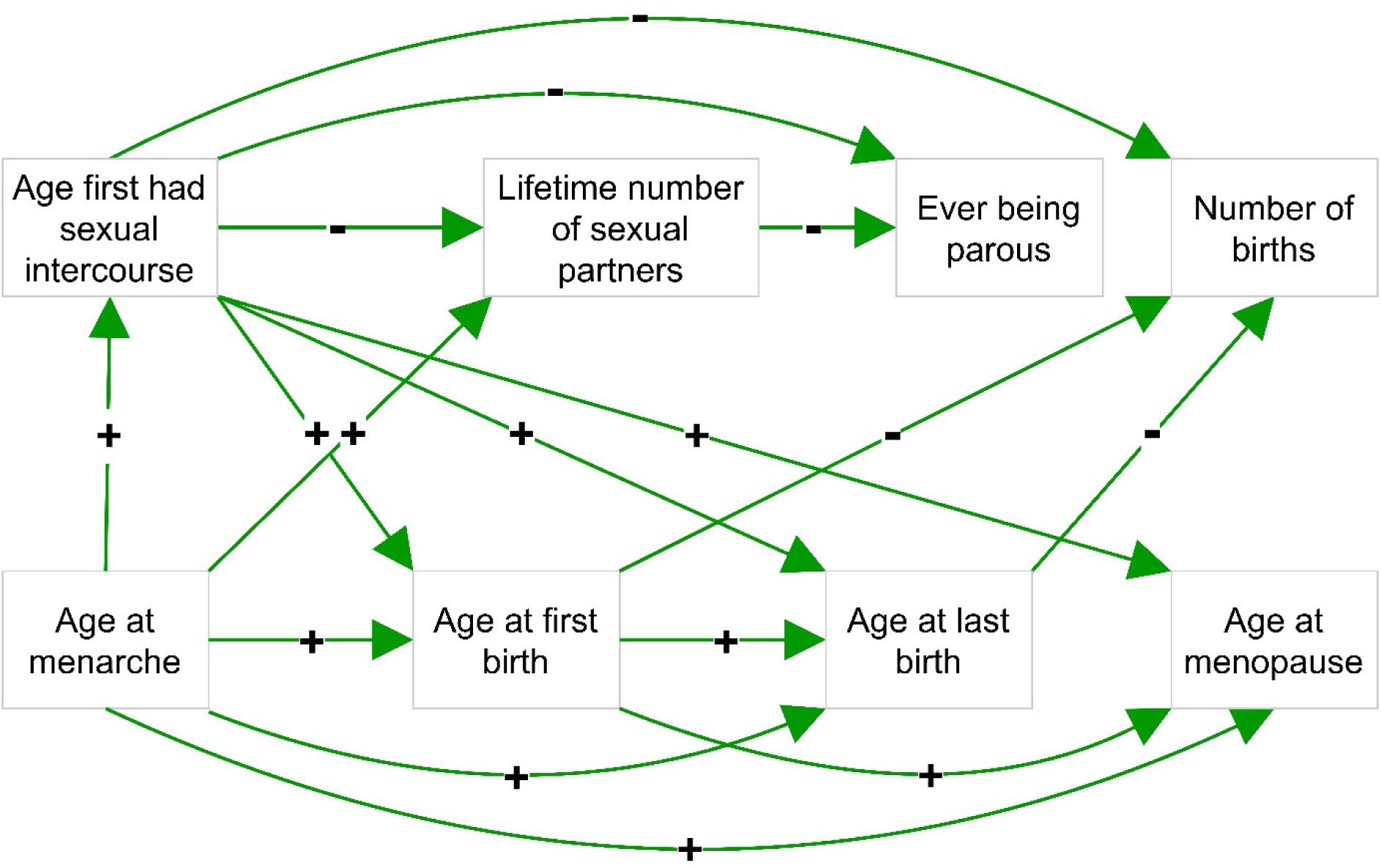
Relationships identified in the primary with evidence of a causal effect. + indicates a positive relationship and – indicates a negative relationship.

### Evaluating the assumptions of MR

#### Negative controls

We found little evidence for an effect of age at menopause on AFS (B=0.03 SD, CI=-1.32×10^−3^, 0.05), of ALB on AAM (B=0.11 SD, CI=-0.12, 0.34), of AFB on AAM (B=0.04 SD, CI=-0.07, 0.16), or of AFS on AAM (B=0.10 SD, CI=-7.03×10^−4^,0.21) (**Table S7**). However, there was strong evidence for an effect of AFB on AFS (B=0.58 SD, CI=0.52, 0.65), suggestive of shared pleiotropy.

#### Heterogeneity

For the relationships identified in the primary analysis, evidence for heterogeneity in the individual SNP effects in the IVW was present across many of the investigated relationships, except for between AFB and ALB, and between ALB and number of births (**Table S8**). Evidence for heterogeneity could indicate the presence of SNP outliers which were investigated using MR PRESSO (see “Pleiotropy”).

#### Pleiotropy

The effects of AAM on AFS, of AFS on AFB and ALB, and of AFB on ALB and menopause and number of births, were consistent across MR Egger, Weighted median, and Weighted mode that test for presence of pleiotropy (**Table S9, Figure S1**).

Effects were less consistent across the additional MR methods between AAM and AFB, ALB, menopause, and lifetime number of sexual partners, as well as between age first had sexual partners and age at menopause, lifetime number of sexual partners, number of births and ever being parous. Furthermore, the effect of AFB on age at menopause, ALB on number of births and lifetime number of sexual partners and ever being parous appeared inconsistent across the different MR methods. In the primary analysis, the only instance where the MR-Egger intercept test revealed evidence for directional pleiotropy was in the relationship between AAM and lifetime number of sexual partners (**Table S10**). We also applied MR-PRESSO to the UK Biobank full overlap GWAS to additionally test for evidence of pleiotropy and correct for outliers (**Table S11**). MR-PRESSO revealed evidence for outliers in almost all tests, other than for the relationships between AFB and ALB. However, after outlier-correction, there was little change in the strength of evidence.

We applied an MR Steiger method to assess whether we had captured the intended causal direction between reproductive factors where the causal direction was unclear. Findings show aggregated instruments have successfully captured the intended causal direction in all cases (**Table S12**). Steiger filtering was also implemented to assess whether there were any individual SNPs that did not capture the intended causal direction, and results are displayed in **Table S13**. Where instruments contained SNPs that did not capture the intended causal direction, MR analysis was then performed excluding those SNPs and the strength of evidence for the causal estimate using the IVW method did not change (**Table S14**).

### Assessing the impact of sample overlap

#### UK Biobank split-sample

In the split-sample GWAS within UK Biobank, between 1 and 101 SNPs were identified at genome-wide significance (p<5×10^−8^) after LD clumping (r^2^ < 0.001 and a distance of 10 000 kb) (**Table S15**). No SNPs were identified at genome-wide significance in relation to ALB and parous status in the GWAS performed on one of the UK Biobank split-samples, therefore the split-sample MR was only conducted once when ALB or ever parous status was the exposure.

Where SNPs were identified in the split-sample analysis, F statistics were above the standard threshold of 10, indicative of strong genetic instruments (**Table S16**). However, there was little overlap in the SNPs which surpassed genome wide significance between sample 1 and sample 2, with 9 SNPs overlapping between samples for AAM and age at menopause but none for the other traits (**Table S16**). A number of the SNPs identified in one of the samples of the split sample GWAS were identified above the significance threshold but remove during LD clumping in the GWAS of the other sample, while other SNPs were just below the significance threshold or appeared not to be associated (**Table S17**).

We performed MR for each relationship twice, i.e., MR of exposure in sample 1 on outcome in sample 2, and MR of exposure in sample 2 on outcome in sample 1. This was with the exception of the MR analyses when ALB and parous status were the exposure, which were assessed only once (**Table S18**). We then meta-analysed findings between both samples, which showed limited evidence of heterogeneity between the causal estimates obtained from the split-sample MRs. Full results of the meta-analysis can be found in **Table S19**.

#### Replication consortia

Inter-relations between the reproductive factors were also investigated using GWAS summary statistics from consortia studies which excluded UK Biobank. 60 SNPs were identified at genome wide significance (p<5×10^−8^) for AAM (ReproGen), and 5 for AFB (SSGAC) (**Table S20**). All F statistics were above the standard threshold of 10, indicative of strong genetic instruments (**Table S20**). Full results of this analysis can be found in **Table S21**. Estimates were consistent when using a larger replication GWAS from ReproGen for age at menopause, although the sample this GWAS was performed in had a large proportion of UK Biobank overlap (**Table S4**).

#### MRlap UK Biobank

MRlap was performed using the reproductive factor GWAS summary statistics for the full UK Biobank sample. This method identified slightly more variants at genome-wide significance (p<5×10^−8^) after LD pruning (10 000kb, r^2^=0.001) compared to the main analysis. Between 11 (ALB and number of births) and 231 (AAM) were identified (**Table S22**). MR estimates were largely similar to the primary analysis, although in some cases effect size were slightly larger, including for the relationship between age at last birth and number of births. Full results of this analysis can be found in **Table S23**.

#### Assessing evidence of causal effects across sensitivity analyses

**Figure 4** illustrates the effects which appear robust across multiple sensitivity analyses. In particular, a later AAM appears to have consistent effects on a later AFB, ALB and AFS. In addition, a later AFB leading to a later ALB, later AFS leading to later AFB, and a later AFS leading to a lower number of lifetime sexual partners, were consistent across all sensitivity analyses. There was no consistent evidence for a causal relationship between AAM and lifetime number of sexual partners across sensitivity analysis and limited evidence between AFS and age at menopause. (**Figure 4**)

**Figure 4.**
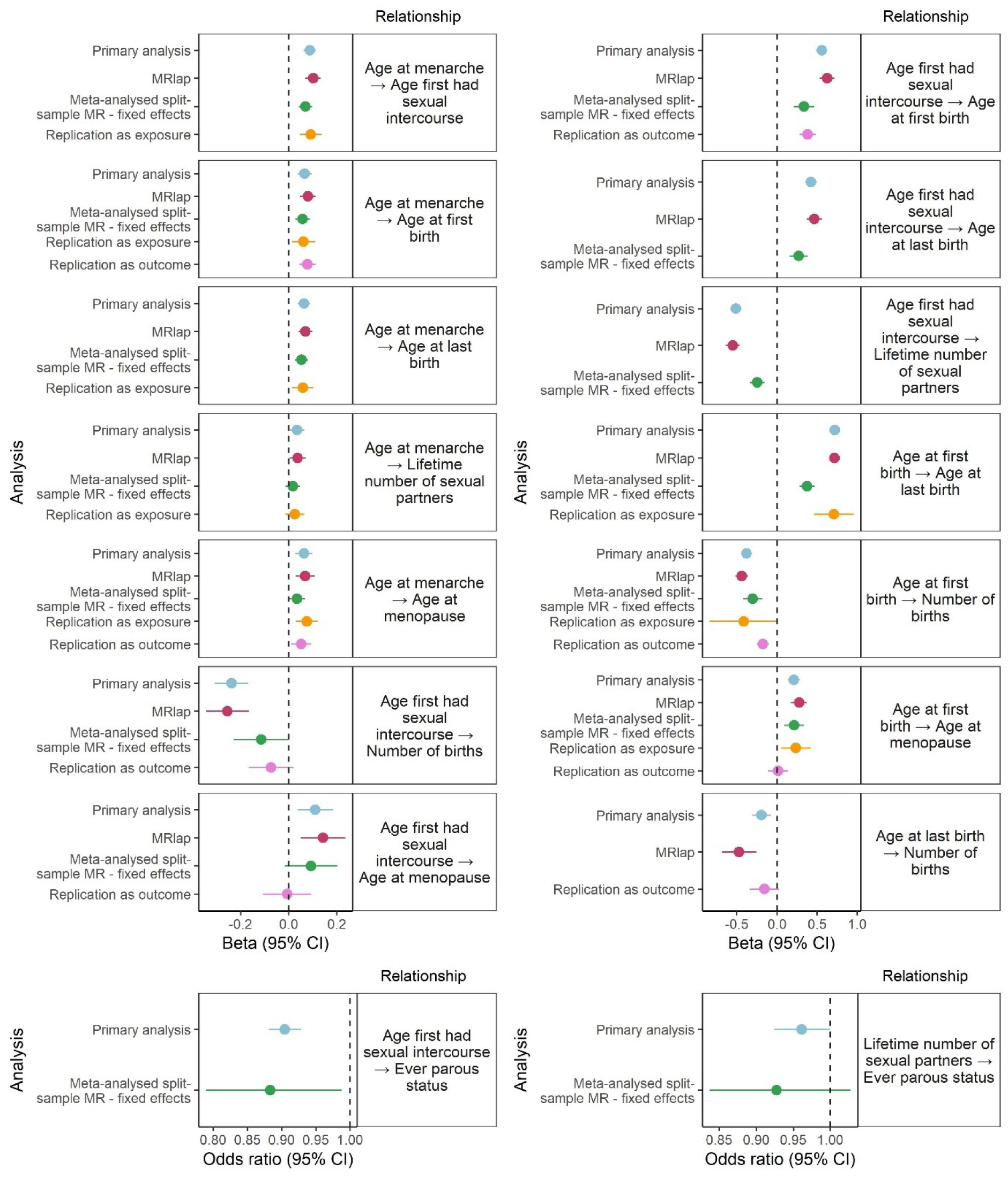
Forest plot showing findings from the primary MR analysis and across the sensitivity MR analyses (IVW MR method). Panels to the right of the plots refer to the relationships investigated, and each analysis is shown on the y axis.

## Discussion

This study provides evidence supporting causal effects of several female reproductive factors on other reproductive traits. We show evidence that earlier reproductive factors including AAM, AFS and AFB have effects on subsequent events and factors, while ever parous status, age at menopause, number of births, ALB and lifetime number of sexual partners appear to have limited effects on other reproductive factors.

We substantiate the genetic correlation between reproductive factors shown in previous studies, while showing additional correlations that have not been previously investigated.^(57,58)^ Our study supports evidence for a positive causal link between AAM and age at menopause,^(11-16,59,60)^ and opposes previous studies that have shown the inverse association,^(17,18)^ or no association.^(19-24)^ Furthermore our findings support one study that found little evidence for an association between AAM and parity.^(26)^ Additionally, we corroborated the findings of previous MR studies that identified a positive causal relationship between AAM and AFB, ALB and age at menopause, and between AFS and ALB.^(59-61)^

Many estimates identified in the primary analysis appear consistent across sensitivity analyses that aim to account for biases. However, some results did not persist in sensitivity analyses checking for robustness to sample overlap and winner’s curse. The split-sample meta-analysed MR shows a weaker magnitude of effect compared to our primary analysis, which may be due to sample size reduction in this sensitivity analysis or bias introduced by sample overlap in the primary analysis. Overall, using replication GWAS studies as the exposure or outcome showed weaker strength of evidence and/or magnitude of effects, although evidence for a causal effect for many relationships assessed was maintained. This may be due to bias introduced by winner’s curse in the primary analysis or smaller sample sizes available for the replication studies. In particular, age at menopause from the ReproGen consortium has a sample size of 69,360, compared to 143,791 in our primary analysis, and where this is used as the outcome, we found little evidence of an effect of reproductive factors on age at menopause. A more recent GWAS of age at menopause conducted by the ReproGen consortium has a much larger sample size (n=201,323), ^(62)^ although more than half of the sample comprise UK Biobank women, meaning a large sample overlap in the MR analysis. Nonetheless, MR estimates using this more recent GWAS revealed similar results compared to the previous smaller GWAS.^(32)^ MRlap revealed almost identical results compared to our primary analysis suggesting sample overlap may not substantially bias estimates.

Pleiotropy may occur when genetic variants have an effect on multiple phenotypes, which can be an issue in MR as the genetic instruments used as a proxy for the exposure can affect the outcome independently of the exposure of interest.^(29,55)^ Therefore, resulting effect estimates may not correctly capture the exposure-outcome relationship of interest. This could be a problem as many of the reproductive factors are genetically correlated, and consequently multiple sensitivity analyses were used to assess whether there was an exclusion restriction assumption violation. We implemented additional MR methods and numerous relationships did not appear to be affected by pleiotropy. However, it is worth considering that a recent study found that using MR Egger on overlapping exposure and outcome samples may induce bias in the direction and magnitude of the confounding.^(43)^ Conversely, other two-sample methods appear to perform similarly in a one-sample MR compared to a two-sample approach in similarly large sample size.^(43)^ Therefore, where the MR Egger estimate deviated from the effects estimated from other MR approaches, caution is warranted in MR Egger interpretation. Where outlier-correction was possible, results were consistent with the primary analysis, with the exception of the effect of lifetime number of sexual partners on ever having children, where there was a complete attenuation of the effect after outlier-correction.

### Mechanisms underlying causal links

We show that an earlier AAM may lead to an earlier AFS and AFB, as well as an earlier AFS leading to an earlier AFB. It is likely that earlier maturation may lead to earlier sexual activity, logically increasing the chance of an earlier pregnancy. In UK Biobank, a proportion of women may have had first had sexual intercourse prior to the introduction of the NHS family planning act 1967 which made contraception readily available through the NHS. This may have strengthened the effect of AFS on AFB in this cohort and findings may not be generalisable to more contemporary studies. We also show that an earlier AFS may lead to a higher number of sexual partners, which may occur due to a longer amount of time to acquire partners if sexual activity commences earlier. Furthermore, we identify that having a higher lifetime number of sexual partners may lead to a lower chance of having children. This may be due to increased prevalence of short term relationships and regularly changing sexual partners,^(63)^ which, as a result, might lead to less chance of starting a family. However, it is worth noting that after excluding outlying variants, the effect between lifetime number of sexual partners and ever parous status attenuated. We present strong evidence for a positive relationship between AFB and ALB. One explanation for this link could be as parents tend to have children in a relatively short period of time, as shown in UK Biobank where the average AFB is 26 years, and ALB is 30 years for women.

The life history theory is another explanation as to why earlier AAM leads to earlier subsequent reproductive events and a likelihood of an increased number of children. This theory distinguishes the allocation of resources into growth and reproductive efforts and categorises “fast” or “slow” life history strategies.^(64,65)^ A “fast” life history strategy exerts more effort towards reproduction: earlier puberty and sexual activity leading to an early AFB, and an increased number of births.^(64,65)^ This is corroborated by our finding that women who experience an earlier AFS, have children earlier and have more children. If woman starts having children earlier, they have more opportunity to conceive again before menopause, which may explain the effect we identify between an earlier AFB and higher number of children. A “fast” life history may lead to an earlier age at menopause as allocating resources towards reproductive efforts earlier in life and towards a higher number of children, which may result in a completing reproduction at a younger age.

There were a number of relationships where we did not find evidence for an effect in our primary analysis. Of note we did not find a causal effect of AAM on number of births and ever parous status. Considering the life history theory, we might have expected to find an inverse effect, suggesting an earlier AAM leads to a high number of births.

Furthermore, we did not find evidence of an effect of ever parous status on lifetime number of sexual partners, number of births on age at last birth. We investigated bidirectional effects between reproductive factors where there wasn’t a clear temporal order and identified no bidirectional effects. Specifically, no effects between age at menopause and age at last birth, lifetime number of sexual partners, number of births and ever parous status, age at last birth and lifetime number of sexual partners, and finally number of births and lifetime number of sexual partners.

Several relationships between reproductive factors separated by many years could be mediated by other intervening reproductive events. For example, we identify effects between AAM and AFS, AFS and AFB, and AAM and AFB, therefore the effect we find between AAM, and AFB may be mediated by AFS. Similarly, we found effects between AFS and AFB, AFB and ALB, and AFS and ALB, which could suggest that an earlier AFS leading to an earlier ALB may be mediated through an earlier AFB. Future investigations could use mediation analyses to further elucidate these relationships.^(66)^

### Implication of findings

When investigating one reproductive factor in relation to a health outcome, our findings might aid in identifying reproductive factors that could confound this relationship. For example, becoming a parent at an earlier age has been identified as a risk factor for depressive symptoms in young adulthood.^(67,68)^ We have presented evidence that AAM has a causal effect on AFB, and previous studies have identified earlier AAM as a risk factor for poor mental health outcomes.^(69,70)^ The evidence presented in this study suggests it would be important to adjust for AAM in an investigation of the effects of AFB on mental health outcomes.

Our work also suggests that reproductive factors might lie on the causal pathway between an earlier reproductive factor and a later outcome. We present evidence for a causal effect between AFB and number of births, and both reproductive factors been identified as a risk factor for cardiovascular disease.^(71)^ An investigation of AFB on risk of cardiovascular disease might want to consider mediation via number of births.

Finally, a number of reproductive factors have been identified as risk factors for breast cancer, including AAM, age at menopause,^(2)^ number of births and AFB.^(3)^ We have presented a number of causal inter-relationships between reproductive factors; therefore, researchers should carefully consider the total impact of reproductive factor variability on chronic diseases such as breast cancer rather than the impact of single reproductive indicators, a multivariable approach could be particularly useful.^(72)^

### Strengths and Limitations

The strengths of the study include the range of reproductive factors investigated using the MR approach; use of large UK Biobank resource and data from other genetic consortia; and, the extent of MR sensitivity analyses to evaluate MR assumptions and address sample overlap. However, this study has a number of limitations.

Firstly, negative control analysis revealed strong evidence of an effect of AFB on AFS, suggesting possible evidence of pleiotropy which has been previously identified for the AFS genetic instrument.^(73)^ As this may reduce the reliability of our results, future work could further assess whether the associations identified for AFS reflect true causal effects.

For some exposures such as ALB, number of births and ever being parous, the number of SNPs used as genetic instruments was limited, meaning we cannot reliably evaluate pleiotropy and heterogeneity in these instances. Increasing the number of SNPs in the genetic instruments for each of these reproductive factors through larger GWAS would be valuable.

Another limitation is the issue of selection bias in UK Biobank. While 9 million individuals were invited to participate in the study, the response rate was 5%. Additionally, the participants in the UK Biobank and replication studies we used were largely restricted to women of European ancestry. These samples are therefore not representative of the entire UK female population and estimates may not be generalisable to women in other ancestry groups. Future work is required to replicate our findings in independent studies and translate the results in women in other ancestry groups.

While the majority of the reproductive factors are likely to be accurately captured through questionnaire (such as AFB, number of births and ALB), other factors such as AAM may not be as reliably recalled ^(74)^. Self-report of lifetime number of sexual partners is also known to be overestimated by some, which could explain the positively skewed distribution we identified.^(75)^ To account for this we performed rank-based inverse normal transformation of this variable.

The split-sample GWAS revealed little overlap between genome wide significant SNPs identified in each sample. While some of these SNPs were identified slightly below the significance threshold between samples, others appeared not to be associated. This suggest that some SNPs may have been identified through spurious associations and may suggest evidence of winner’s curse.

## Conclusion

In conclusion, we present evidence of inter-relationships between reproductive factors. In particular, we find strong evidence of an effect of AAM, AFS and AFB on subsequent reproductive events and factors. Future work should consider the inter-relationships between reproductive factors when assessing reproductive risk on disease outcomes.

## Data Availability

The availability of all data analysed in this study has been referenced throughout the manuscript and supplementary materials.

https://www.reprogen.org/

https://www.thessgac.org/

## Acknowledgements

This research was conducted using the UK Biobank Resource under application number 6326. We thank the participants and researchers from the UK Biobank who contributed or collected data. This work was carried out using the computational facilities of the Advanced Computing Research Centre, University of Bristol - http://www.bris.ac.uk/acrc/.

