## Supplementary Note for "The relationships between women’s reproductive factors: a Mendelian randomization analysis"

### **Supplementary Figures**

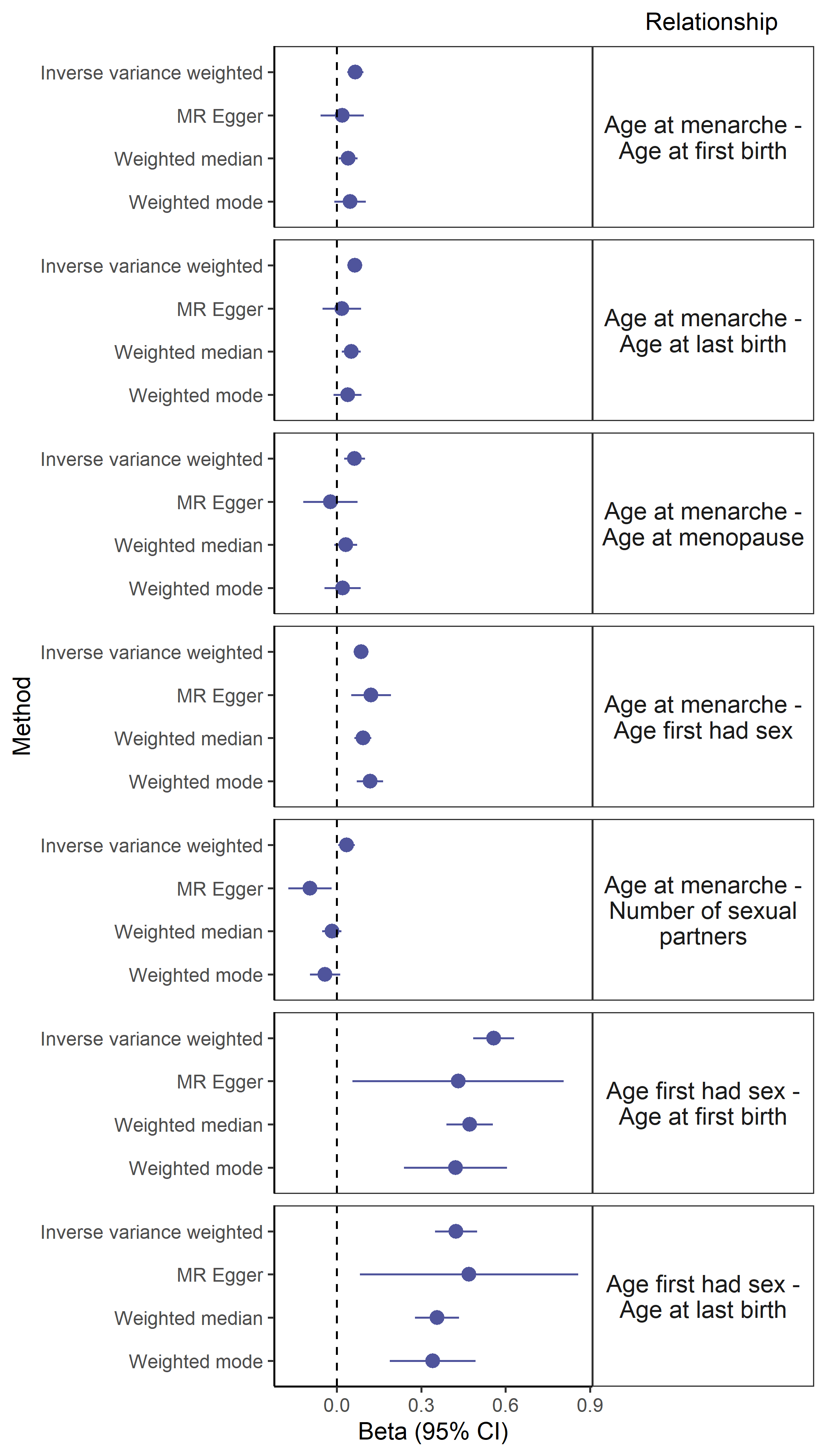

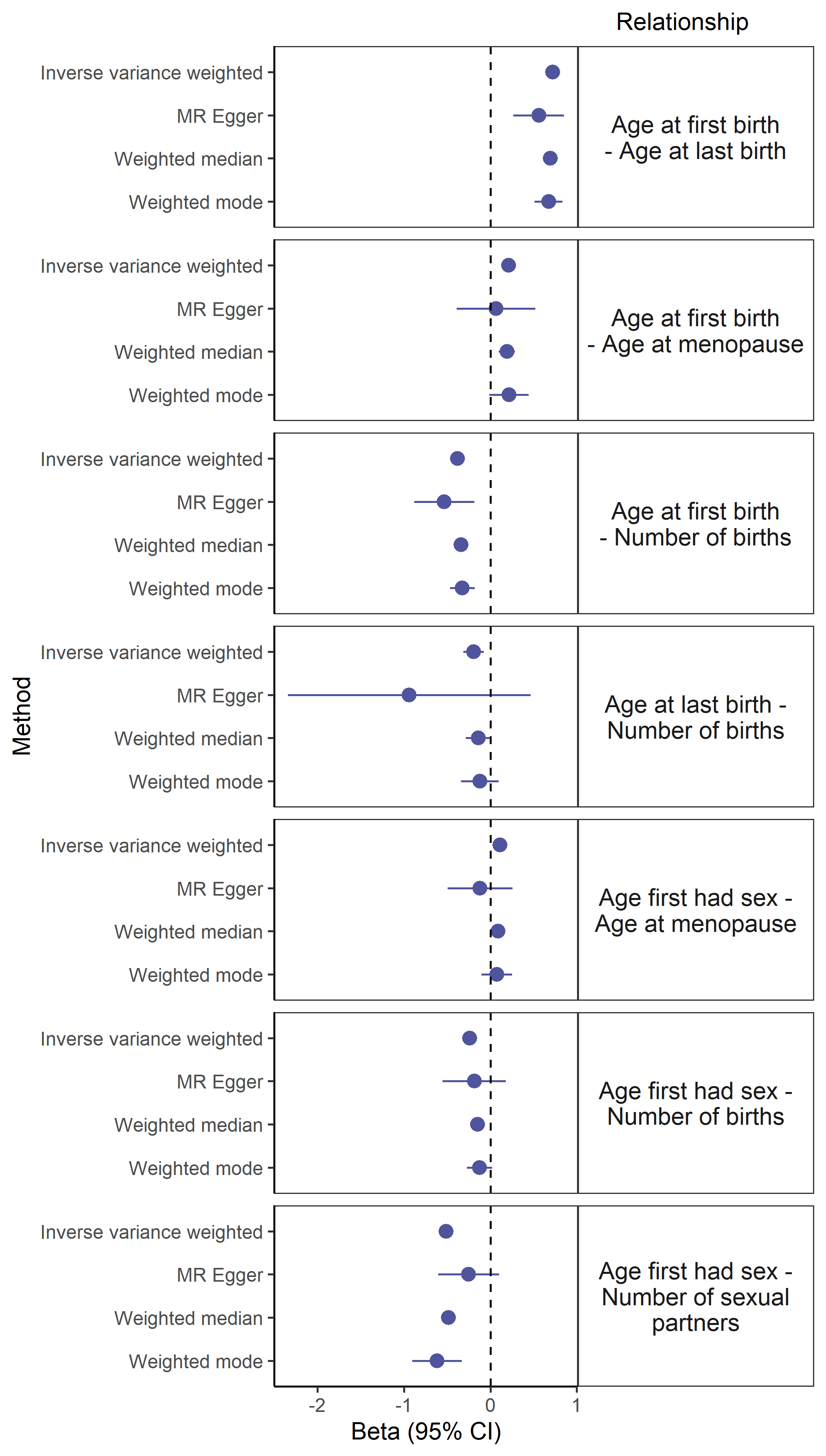

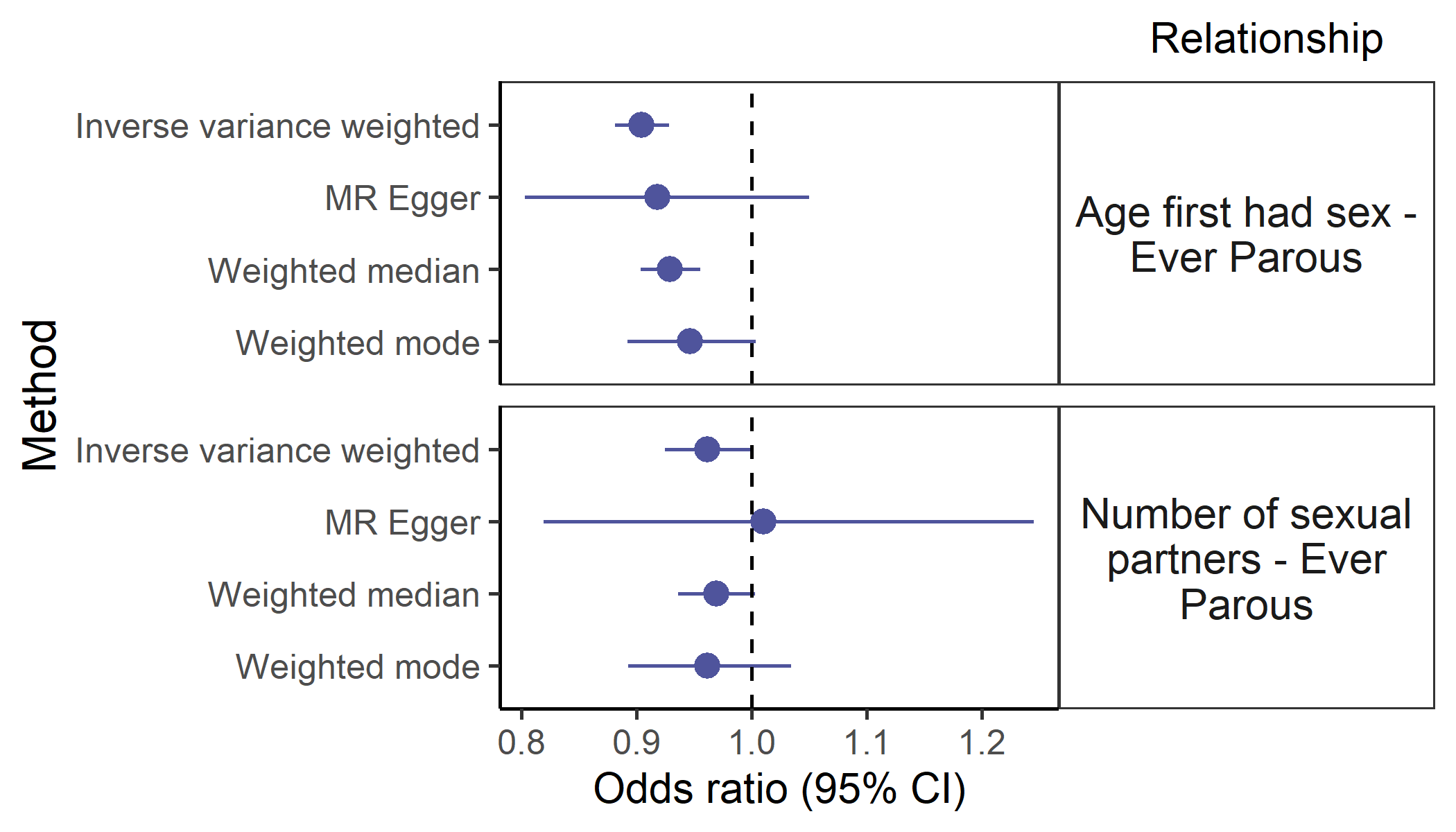

**A**

**B**

**C**

**D**

**E**

**F**

**G**

**O**

**P**

**H**

**I**

**J**

**K**

**L**

**M**

**N**

Figure S1 Forest plots showing effect estimates of additional MR methods for relationships identified in the primary MR analysis. Panels A-P refer to the relationships assessed using MR, and MR methods used is shown on the y axis.

### **Supplementary note**

#### **Methods**

##### **UK Biobank reproductive factors**

Age at menarche was derived from the question: "How old were you when your periods started?". Age at menopause was derived from the question "How old were you when your periods stopped?". To identify the number of live birth births, women were asked "How many children have you given birth to? (Please include live births only)". A binary measure of parous status, coded as “0” or “1”, where women who had or had not given birth, was derived from the measure of number of live births. Women who indicated that they had given birth to one child were asked "How old were you when you had your child?". Women who indicated that they had given birth to more than one child were asked "How old were you when you had your FIRST child?" and "How old were you when you had your LAST child?". To derive age at first live birth and age at last live birth, responses from primiparous and multiparous women were combined. Women were asked also asked, "What was your age when you first had sexual intercourse? (Sexual intercourse includes vaginal, oral or anal intercourse)" and "About how many sexual partners have you had in your lifetime?", collected from all participants except those who indicated they never had had sexual intercourse. Responses to these questions were used to derive age first had sexual intercourse and lifetime number of sexual partners, individuals who had not had sexual intercourse were coded as “0” in the number of sexual partners analysis. We performed rank-based inverse normal transformation on lifetime number of sexual partners given the skewed distribution of this variable, all other RFs were untransformed.

#### **Evaluating MR assumptions**

##### Pleiotropy

We performed additional MR methods: Weighted mode,^(1)^ Weighted median,^(2)^ and MR Egger, to assess evidence of pleiotropy.^(3)^ MR Egger allows for pleiotropic SNPs assuming pleiotropy satisfies the Instrument Strength Independent of Direct Effect (InSIDE) assumption which states that the SNP-exposure association is not correlated with path from the instrument to the outcome that is independent of the exposure, unbalanced pleiotropy violates the InSIDE assumption.^(4,5)^ The intercept and 95% confidence interval of the MR-Egger regression line was used to determine directional pleiotropy using the TwoSampleMR package.^(6)^

We also applied the R function MR-PRESSO (Mendelian Randomisation Pleiotropy RESidual Sum and Outlier) to evaluate the MR results by identifying and correcting for potential outliers (p <0.05)^(7)^. MR-PRESSO allows the evaluation of genetic pleiotropy in the MR model, performs outlier removal, and performs MR again without outliers to reduce bias in MR estimates.

##### Steiger filtering for bidirectional relationships.

For those reproductive factors investigated bi-directionally where temporal ordering was not clear, we performed the MR Steiger test and Steiger filtering to assess whether the hypothesized causal directional of the relationship was correct for each genetic instrument. ^(8)^ Both were performed using the UK Biobank GWAS summary statistics where the exposure and outcome sample overlapped, using the “TwoSampleMR” package, the MR Steiger test using the function “mr_steiger()”, and Steiger filtering using “steiger_filtering()”.

#### **Assessing the impact of sample overlap**

*MRlap*

MRlap is a recently developed MR method which can account and correct for bias arising from sample overlap, weak instruments and winner’s curse.^(9)^ As previously described bias introduced by sample overlap between the exposure and outcome samples could lead to an overestimation of estimates.^(10)^ MRlap combats sample overlap, for example, by reducing an observed 15% overestimation for fully overlapping samples to a 5% overestimation. Instrument strength is determined by the magnitude and precision of association of the genetic instrument and can be assessed using R^2^ and F statistic, instruments with a F statistic over 10 are considered sufficient. In one-sample MR weak instruments bias estimates towards the confounded exposure-outcome association whereas in two-sample MR where samples do not overlap the bias is expected to be towards the null.^(11,12)^

MRlap using cross-trait LDSC to approximate the sample overlap and both the exposure and the outcome should be continuous traits using this method, as the correction for biases cannot account for a different degree of overlap for cases/controls in case of binary traits.^(9)^ Therefore, MRlap was performed using the UK Biobank GWAS summary statistics for reproductive factors where both the exposure and outcome were continuous i.e., excluding relationships involving ever parous status.
